# Current sampling and sequencing biases of Lassa mammarenavirus limit inference from phylogeography and molecular epidemiology in Lassa Fever endemic regions

**DOI:** 10.1101/2023.06.20.23291686

**Authors:** Liã Bárbara Arruda, Hayley Beth Free, David Simons, Rashid Ansumana, Linzy Elton, Najmul Haider, Isobella Honeyborne, Danny Asogun, Timothy D McHugh, Francine Ntoumi, Alimuddin Zumla, Richard Kock

**Author notes:** School of Life Sciences, Faculty of Natural Sciences, Keele University, Staffordshire, United Kingdom. Both authors contributed equality to this work (Hayley Beth Free, David Simons).

## Abstract

Lassa fever (LF) is a potentially lethal viral haemorrhagic infection of humans caused by *Lassa mammarenavirus* (LASV). It is an important endemic zoonotic disease in West Africa with growing evidence for increasing frequency and sizes of outbreaks. Phylogeographic and molecular epidemiology methods have projected expansion of the Lassa fever endemic zone in the context of future global change. The Natal multimammate mouse (*Mastomys natalensis*) is the predominant LASV reservoir, with few studies investigating the role of other animal species. To explore host sequencing biases, all LASV nucleotide sequences and associated metadata available on GenBank (n = 2,298) were retrieved. Most data originated from Nigeria (54%), Guinea (20%) and Sierra Leone (14%). Data from non-human hosts (n = 703) were limited and only 69 sequences encompassed complete genes. We found a strong positive correlation between the number of confirmed human cases and sequences at the country level (*r* = 0.93 (95% Confidence Interval = 0.71 - 0.98), *p* < 0.001) but no correlation exists between confirmed cases and the number of available rodent sequences (*r* = -0.019 (95% C.I. -0.71 - 0.69), *p =* 0.96). Spatial modelling of sequencing effort highlighted current biases in locations of available sequences, with increased effort observed in Southern Guinea and Southern Nigeria. Phylogenetic analyses showed geographic clustering of LASV lineages, suggestive of isolated events of human-to-rodent transmission and the emergence of currently circulating strains of LASV from the year 1498 in Nigeria. Overall, the current study highlights significant geographic limitations in LASV surveillance, particularly, in non-human hosts. Further investigation of the non-human reservoir of LASV, alongside expanded surveillance, are required for precise characterisation of the emergence and dispersal of LASV. Accurate surveillance of LASV circulation in non-human hosts is vital to guide early detection and initiation of public health interventions for future Lassa fever outbreaks.

## 1 Introduction

Lassa fever (LF) is a lethal zoonotic viral haemorrhagic disease of humans, caused by *Lassa mammarenavirus* (LASV). It causes an estimated 900,000 annual human infections and several thousand deaths in West Africa annually (1,2). The WHO assigns LASV endemicity to eight West African countries: Benin, Ghana, Guinea, Liberia, Mali, Sierra Leone, Togo and Nigeria (S1 Fig) (3). LASV is a bisegmented ssRNA-virus of the family Arenaviridae (4,5). Based on the genomic analysis of the large (L) and small segments (S) LASV has been classified into seven lineages which demonstrate spatial segregation across the endemic range (6). The high nucleotide variability (25-32%) of these lineages introduces complexity into assays to detect LASV infection.

Epidemiological data on LF is limited and constrained by current testing and reporting in the endemic region, making accurate estimates of its true burden challenging (7). Many individuals infected with LASV do not seek healthcare with up to 80% of infections assumed asymptomatic or presenting as mild illness (8). Estimates based on longitudinal serological surveys in Sierra Leone in the early 1980’s indicated that 100,000 to 300,000 infections of LF occurred annually in West Africa, with more recent estimates being up to 900,000 infections (2,8). Identification of symptomatic cases is further confounded by overlapping symptoms with other diseases (e.g., malaria) and lack of available diagnostic methods (1,9–11). Access to diagnostic tests varies spatially, increased availability at centers of excellence in LF treatment and research such as the Irrua Specialist Teaching Hospital, Nigeria and Kenema General Hospital, Sierra Leone results in a spatial bias of reported cases from these locations. Phylogenetic analysis and molecular dating of sequence clinical and research samples suggest a westward route of dispersal of LASV lineages, from the most recent common ancestor in Nigeria. (12–18). These estimates have been used to project the potential for Lassa Fever to extend beyond the current endemic zone (19).

The Natal multimammate mouse (*Mastomys natalensis*) is the primary reservoir of LASV, however, 11 other rodent species have been found to be acutely infected or have seropositivity to LASV including; *Mastomys erythroleucus, Hylomyscus pamfi, Mus baoulei* and *Rattus rattus* (15,20–24). Humans become infected with LASV upon contact with or inhalation of excretions from the rodent species (12,25). Although human-to-human transmission has been reported – typically associated with nosocomial outbreaks – these are rare events when compared with spillover from rodent hosts (26). We performed a study of LASV nucleotide sequences available from the National Centre for Biotechnology Information (NCBI) GenBank, using associated metadata to spatially model sequencing effort, adjusted for the number of suspected and confirmed human LF cases to determine potential biases in locations of available sequences or significant geographic limitations in LASV surveillance, particularly, in non-human hosts.

## 2 Methods

### 2.1 Data Collection and Processing

LASV nucleotide and protein sequences were obtained from the NCBI GenBank (27). The search query run on 24 Sep 2021 was for “Lassa mammarenavirus” in the organism field of the NCBI nucleotide dataset. Data were obtained using the NCBI Entrez API with analysis conducted using the “genbankr” package within the R statistical programming language (27– 29). Associated citations were manually retrieved to identify missing metadata for sequences including hosts and geographic location of samples. Sequences with large portions (10% missing compared to reference sequences, NC_004296.1 and NC_004297.1 for S and L segments respectively) of missing nucleotide data on the L- or S-segment or lacking associated metadata (collection year, host species, country, and geographical region of sampling) were excluded from phylogenetic analysis. Nucleotide sequences were aligned using the ‘map to reference’ tool on Geneious Prime 20201.2. Alignment, visual inspection and manual editing were performed, and entries that contained >100 continuous ambiguous nucleotide calls were excluded (S1 Data).

### 2.2 Sequencing Bias

First, we compared the number of cases reported from countries between 2008-2023 with the number of samples contained in GenBank to summarise the correlation between reported human cases and availability of sequences. We then compared the proportion of human to non-human derived sequences within countries.

To understand the bias of sequenced samples at a sub-national level the origin of a sequenced sample was geocoded using the Google Geocoding API using the “ggmap” package (30). Sequence locations were associated with level-1 administrative regions and data were separated into human and rodent sources of samples to visualise the spatial heterogeneity of sampling. To measure sampling effort bias, the number of samples obtained within a level-1 administrative region was associated with the centroid of the region. The number of confirmed LF clinical cases reported from these regions in the previous 15 years was obtained (S2 Data). The number of cases within a region was divided by the human population count to produce the number of confirmed cases per 100,000 individuals. The number of sequences was used as the response variable in a spatial Generalised Additive Model, with geographic coordinates and cases per 100,000 individuals used as covariates. This model was constructed using the “mgcv” package (31).

### 2.3 Phylogenetic Analysis

Phylogenetic analysis was undertaken through Bayesian Markov Chain Monte Carlo (MCMC) method using BEAST.v1.10.4 (32). In BEAUTi, the parameters were a substitution model as a generalised time reversible plus gamma site heterogeneity, with codon partition positions 1, 2, 3. A strict clock and a coalescent tree prior with a constant size population was used. Each analysis consisted of 20 million MCMC steps and trees were sampled every 20,000 generations. Sample collection dates from the metadata were used as tip dates to fit to a molecular clock, and country of sample collection was incorporated as a discrete state (16,33). To assess the log files of the output TRACER.v.1.7.1 was used. Maximum-clade credibility trees were generated through TreeAnnotator v1.8.4 and visualised in FigTree.v1.4.4 (34).

## 3 Results

### 3.1 Compiled Dataset

The initial dataset comprised 2,298 records (from samples obtained 1969-2019), including nucleotide sequences and associated metadata. Incomplete gene sequences and sequences lacking metadata information (n = 1,045) were removed from phylogenetic analyses. Therefore, 680 sequences of complete S segment and 573 sequences of partial L segment (L protein only) were used. Accession numbers of included and excluded sequences are available in S1 Data.

### 3.2 Descriptive Analysis

Year of collection was available for 2,108 records, with the oldest sequence dating from 1969 and latest from 2019. Among these records, most sequences (n = 1,936, 92%) have been obtained since 2008. Human-derived LASV sequences comprised most of the available records (67%), other host species include *Mastomys natalensis* (29%) and *Mastomys spp*. (3%), while *Mastomys erythroleucus* (n = 18), *Mus baoulei* (n = 9) and *Hylomyscus pamfi* (n = 10) represent < 1% each. The species sampled was not documented in 107 records. Country of collection was available for 2,238 records. Most sequences were produced from samples collected in Nigeria (54%), followed by Guinea (20%), Sierra Leone (14%), Liberia (4%) and Cote d’Ivoire (3%) with the remainder obtained from, Benin, Ghana, Mali and Togo (Fig 1).

**Figure 1.**
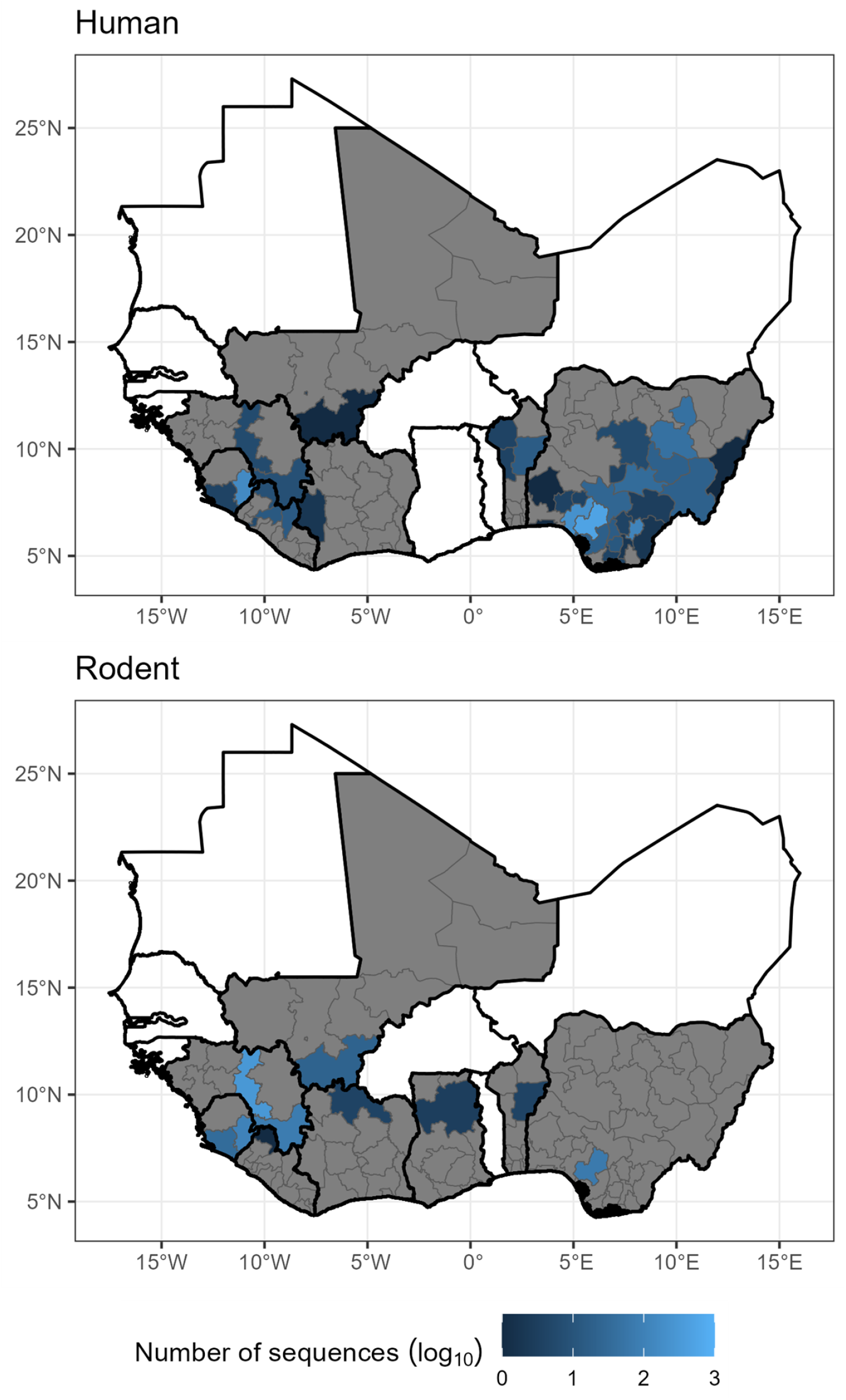
The number of sequences, shown on a log_10_ scale, retrieved from NCBI GenBank with associated regional sampling location and host for human samples (top, n = 1,328) and rodent samples (bottom, n = 527). Grey regions represent level-1 administrative areas with no sequences within countries that have at least one available sequence. White countries are West African countries with no available LASV sequences. See S1 Fig for country names. Shapefiles for basemap layer obtained from GADM 4.0.2 (35)

Sequences for human derived samples with regional location data (n = 1328, 63%) were clustered in Edo State, Nigeria (n = 519, 39%), Ondo State, Nigeria (n = 220, 17%) and Eastern Province, Sierra Leone (n = 159, 12%) with 430 samples from the remaining endemic regions. Sequences from rodent samples with regional location data (n = 527, 25%) were most commonly obtained from Faranah, Guinea (n = 210, 39%) and Eastern Province, Sierra Leone (n = 107, 20%) with 210 samples from the regions.

### 3.3 Sequencing bias

We observed a strong positive correlation between the number of confirmed human cases between 2008-2023 and the number of GenBank deposited sequences at country level (*r*(degrees of freedom = 7) = 0.93 (95% Confidence Interval = 0.71-0.98), *p* < 0.001). When analysed by species source no correlation was observed with the number of confirmed cases and the number of available rodent sequences was observed (*r*(6) = -0.019 (95% C.I. -0.71-0.69), *p =* 0.96).

When combining both human and rodent-derived samples at the regional level to explore spatial sampling biases, we found that sequencing effort is greatest in Southwest Nigeria, centred over Edo State and the Faranah and Nzérékoré regions of Guinea, Eastern Province of Sierra Leone and Nimba district of Liberia (Fig 2). There was a positive, non-linear association between the rate of confirmed human cases with the number of available rodent and human derived LASV sequences at regional level (deviance explained = 14%, estimated degrees of freedom = 2.29, p < 0.001).

**Figure 2.**
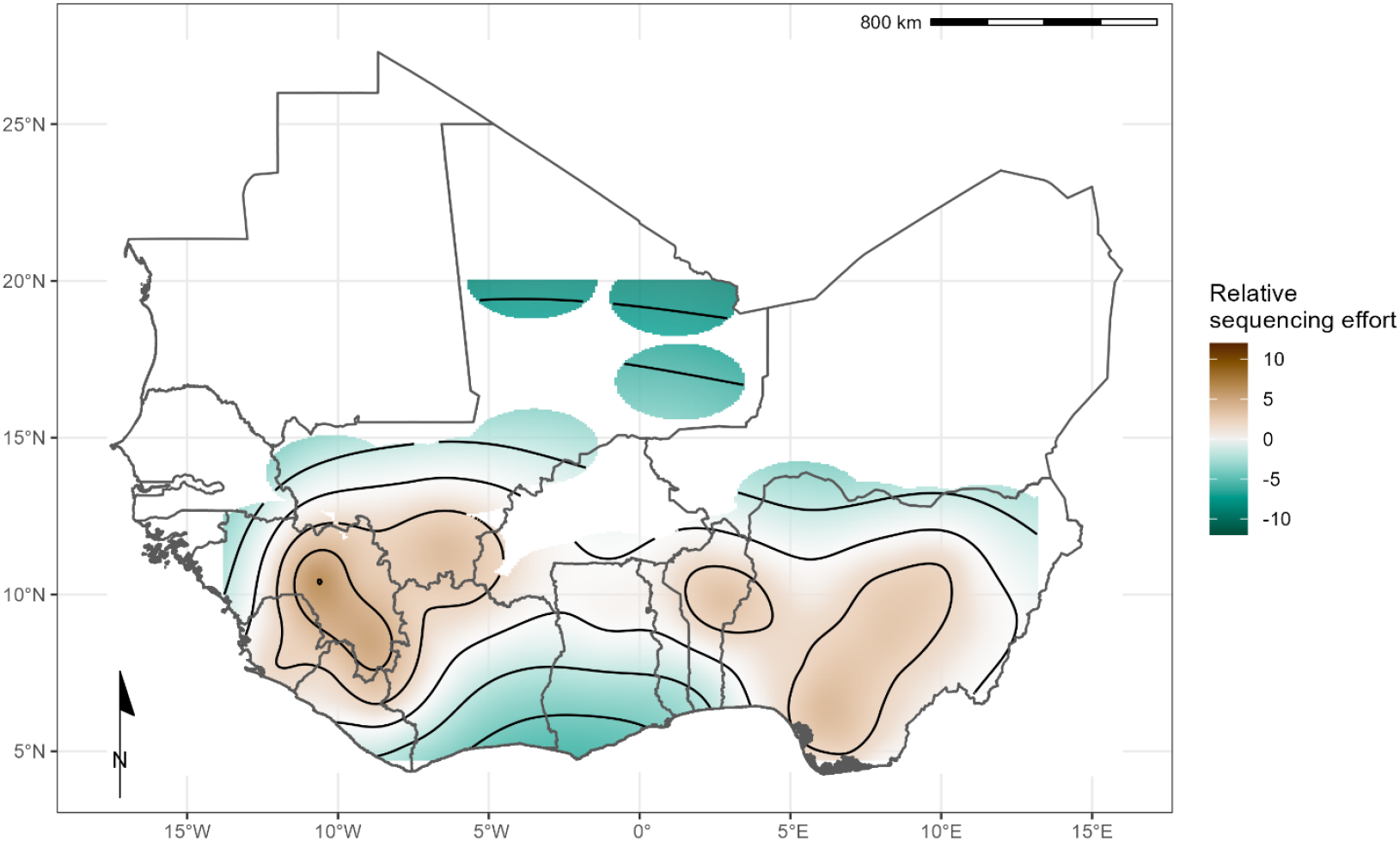
Modelled relative sequencing effort derived from both human and rodent samples. Greatest sequencing effort coincides with areas where sampling in humans (Edo, Nigeria and Kenema, Sierra Leone) and rodents (Faranah, Guinea) have historically been focussed. Shapefiles for basemap layer obtained from GADM 4.0.2 (35)

### 3.4 Phylogenetic Analysis

Sequences for each segment of LASV showed clustering according to previously documented lineages I-VII alongside geographical clustering with lineages I-III and VI present in Nigeria, lV in Liberia, Guinea and Sierra Leone, V in Mali and VII in Togo (S2 Fig). In this analysis only L segment sequences of lineage V from Cote d’Ivoire were included due to quality control exclusion criteria. The phylogeny of the L segment indicates an older emergence of LASV in the human population, with the most recent common ancestor (MRCA) predicted in the year 828 in Nigeria, inference based on the S segment indicates the emergence in the year 1350 (Table 1).

**Table 1.** The most recent common ancestor (MRCA) stratified by host and country of collection of *Lassa mammarenavirus* (LASV) S and L segments. Samples were collected between 1969-2018.

| Host species | Country | S segment MRCA | L segment MRCA |
| --- | --- | --- | --- |
| *Homo sapiens* (n=1181) | Benin | 1995 | 1989 |
|  | Guinea | 1895 | 1871 |
|  | Liberia | 1895 | 1627 |
|  | Nigeria | 1681 | 1498 |
|  | Sierra Leone | 1901 | 1874 |
|  | Togo | 2016 | 2014 |
| *Hylomyscus pamfi* (n=2) | Nigeria | 1681 | 1498 |
| *Mastomys erythroleucus* (n=18) | Guinea | 1975 | 2010 |
|  | Nigeria | 2008 | 2006 |
| *Mastomys natalensis* (n=36) | Guinea | 1938 | 1997 |
|  | Mali | 1951 | 2007 |
|  | Sierra Leone | 1909 | 1979 |

There was a lack of sequence information from lineage I and VI, however, phylogeny suggests these lineages are basal to others in Nigeria (S2 Fig). Lineage VII in Togo is most closely related to Nigerian isolates and potentially diverged between 500-900 years ago. The divergence of lineage III and IV is predicted to have occurred between the years 1332-1551. Introduction to countries west of Nigeria appears to be by dispersal initially to Liberia, followed by Guinea in the 1700s, followed by Sierra Leone and Mali approximately 100 years later. A lack of full segment sequences from lineage V limits calculation of divergence from the most recent common ancestor from lineage IV (approximately 200 years).

## 4 Discussion

There are several important aspects of our study and findings. First, we studied a comprehensive dataset of publicly available full-segment LASV sequences, spanning West Africa and host species, to inform our understanding of the phylogeny of LASV dispersal. Second, we identified substantial variability in the origin of available sequences and completeness of records. Third, we showed strong geographic clustering among lineages supporting prior hypotheses of radiation from both Nigeria and a subsequent introduction into Liberia (19). Fourth, the synthesis of available metadata highlights important gaps in currently available data, including spatial bias in the sequencing of samples and suggests this should be used to inform the design of epidemiological programmes going forward.

Our analyses of 2,298 LASV sequences obtained from GenBank highlights the spatial biases in the availability of sequence data that may limit our understanding of the current and historic dispersal of LASV lineages in West Africa. First, sequence data was typically obtained from three of the eight endemic countries: Nigeria, Guinea and Sierra Leone. We found a strong associated between the number of reported human cases and number of available sequences. When stratifying by host species this trend did not remain with rodent derived samples showing no association with the number of human cases indicating important under-sampling in high human cases regions and relatively high sampling in locations with low numbers of human cases. This is potentially an important source of bias when attempting to infer phylogeography within the reservoir host of this zoonotic pathogen. Sequence data from other countries, and more regions within them, across West Africa are required to increase confidence in the timelines of the currently inferred westward expansion. Greater focus needs to be placed on acquiring sequences from the rodent host to understand viral genetic diversity within the primary reservoir species. Comparing rodent derived sequences with those acquired from spillover into human populations may also allow identification of genetic drivers of transmission (36).

The overrepresentation of data from these three countries has been mapped as relative sequencing effort to identify regions where increased LASV sequencing are required to counteract current sequencing biases. Second, geographic clustering of LASV lineages, suggest isolated events of human-to-rodent transmission and the emergence of LASV dating from 1498 in Nigeria. Similarly, Olayemi *et al*. report evidence of earlier emergence of the virus in humans than in rodents in Nigeria (16). Comparatively limited data from non-human hosts with limited genome coverage, (69/703 sequences encompassed complete genes) produce important uncertainty around the observation of human-to-rodent transmission. Taken together, this data highlight limited surveillance among animal species, necessitating further investments in data acquisition and sharing to accurately define the spatiotemporal expansion of LASV in West Africa.

The phylogenetic analysis of LASV stratified by host species supports spatial evolution, in addition to intra-host viral evolution (S2 Fig). For instance, LASV sequences from *M. erytholeucus* sampled in Nigeria and Guinea clustered within lineages III and IV, respectively. Interestingly, these isolates appear to occur after the emergence of the most recent common ancestor virus circulating among humans and *M. natalensis* in these countries (Table 1), suggesting introduction of LASV into *M. erythroleucus* populations was a consequence of pathogen circulation in human and *M. natalensis* populations. Sequences from *M. natalensis* in Sierra Leone exhibit minimal clustering, and were interspersed with sequences from humans, potentially representing isolated events of pathogen introduction into human populations with spillback into commensal rodent populations (i.e., reverse zoonosis). The most recent common ancestor of LASV sequences from *M. natalensis* in Sierra Leone suggest a later emergence of the virus in this country. Our findings corroborate those of Olayemi et al., that within Sierra Leone LASV appears to have emerged in human hosts before rodents (16). However, this data must be caveated by the limited information from rodent species in these locations.

There is a lower coverage of rodent-derived LASV sequences, with those from the primary reservoir *M. natalensis* forming fewer than one-third of all sequences (n = 642, 28%), with substantially lower sampling of other possible rodent hosts, including other *Mastomys* species. Rodent sampling has not increased at the same rate as human samples despite increased sampling effort since 2008 (15,22,37). There is substantial heterogeneity in the locations in which rodent and human samples are available. For example, a relatively high number of rodent samples (n = 429) have been obtained from Guinea while few human sequences (n = 20) are available from these locations. The inverse is true of Nigeria where most human derived sequences are obtained (n = 1,147) but only 85 rodent sequences are available, and all of these from a single state (Edo, Nigeria). The number of suspected and reported cases was found to be positively but non-linearly associated with the number of available sequences. This is suggestive of a consolidation of research and focus of sampling in areas historically with high numbers of human cases but has led to a paucity of sequences from elsewhere in the endemic region. The limited number of full segment sequences from rodents, from few geographic locations, limits our understanding of viral radiation in rodent hosts, particularly from species which are not considered the primary reservoir, e.g., *H. pamfi*. The most recent common ancestor for the viral sequence obtained from *H. pamfi* is estimated to be in the late 1600s, it is therefore possible lineage VI and/or *H. pamfi* as a reservoir of LASV has gone undetected due to lack of sufficient sampling (15).

Interpreting available LASV sequences is challenging for several reasons. A large proportion of available sequences (70%) have been obtained within Lassa fever research programs, representing spatial ascertainment bias (38–40). In addition to these spatial biases’ temporal biases are apparent. Since 2016 there has been a substantial increase in the number of LASV sequences available in NCBI GenBank, reflecting increasing research effort, availability of sequencing platforms and increased data collection during Lassa fever epidemics, such as in the 2018 Nigeria Lassa fever outbreak (41–43). There are notably fewer recorded sequences of LASV from Benin, Togo, and Ghana, suggesting a potential a gap in surveillance and research capacity in these locations or a lack of circulating LASV, despite several reported outbreaks (44–46). Phylogenetic analysis on 60% of our initial dataset, following removal of sequences due to incompleteness or missing geographic and year of collection information (n = 1,045) demonstrated geographic clustering of LASV lineages, supporting prior analyses (14–16,33,44,47–49). Increased data availability from Nigeria following increased LASV surveillance allowed regional analysis of phylogeny for lineages II and III supporting previous findings of expansion of these lineages from North-East Nigeria to the South-West of the country (13,50,51).

A substantial number (n = 869) of the sequences retrieved corresponded to short fragments (< 1 Kb) probably derived from PCR products used for diagnostic purposes rather than for viral genomic surveillance. LASV is a segmented virus, and it was not possible to identify complete genome sequences since both S and L segments are reported separately on the sequence’s repository. The molecular clock analyses from L protein indicated an earlier emergence of LASV when compared to S segment analysis (828 and 1350 respectively), potentially because the viral RNA polymerase (L protein) is less affected by selective pressure than the S segment (12,47,52).

Despite these challenges, this study has synthesised currently available data on LASV sequences to investigate the location and period of sampling to reconstruct the dispersal of viral lineages across the endemic region. Despite the regionalisation of LF being driven by rodent-to-human transmission, there remains scarce LASV genomic data from non-human hosts. We have mapped the locations of relative under sampling to guide targeted efforts to counteract biases in currently available data for both rodent and human derived sequences. Expanded sampling of LASV from animal species within the endemic region will improve our current understanding of LASV evolution and ecology and improve confidence in current estimates of westward expansion of Lassa fever in humans. Further understanding of the viral evolution dynamics of LASV and spatial expansion of current lineages will be vital to ensure adequate diagnostic tools are available to respond to the expected sporadic outbreaks of Lassa Fever across the region.

## Data Availability

All data used in these analyses are publicly available from GenBank. The accession numbers of records used are available as supplementary material. Code to reproduce the metadata analyses are available as an archived Git release on Zenodo (https://doi.org/10.5281/zenodo.6340162)

## Supplementary material

**S1 Data. GenBank accession number of analysed sequences**. This dataset includes available data about host, country, region, year, sequence length, genome segment (L or S) and predicted MRCA.

**S2 Data. Dataset on confirmed Lassa fever cases**. This presents the number of confirmed cases of Lassa fever reported from countries between 2008 and 2023 at a subnational level that were used to calculate the number of cases per 100,000 people. References for the reports used to produce this dataset are included.

**S1 Figure. Map of West Africa**. displays a map of West Africa with country names for reference with Fig 1 and Fig 2. Shapefiles for mapping obtained from GADM 4.0.2 (35)

**S2 Figure. Time-calibrated phylogeny for both the small segment (S) and large segment (L) from included LASV sequences**.

## Author contributions

Conceptualisation: DS and LBA; Methodology: HF, DS, DA and LBA; Formal Analyses: HF, DS and LBA; Investigation: HF, DS and LBA; Supervision: LBA; Data Curation: HF and DS; Writing – original draft preparation: HF, DS, LBA; Writing – Review and Editing: IH, LE, NH, RA, RK, FN, DA, AZ and TMcH; Funding acquisition: AZ and FN.

## Acknowledgements

Linzy Elton, Timothy D McHugh, Francine Ntoumi, and Alimuddin Zumla acknowledge support from EDCTP-Central Africa and East African Clinical Research Networks (CANTAM-3, EACCR-3). Sir Zumla is an NIHR Senior Investigator, a Mahathir Science Award, Sir Patrick Manson Medal and EU-EDCTP Pascoal Mocumbi Prize laureate. Liã Bárbara Arruda, David Simons, Rashid Ansumana, Linzy Elton, Najmul Haider, Isobella Honeyborne, Danny Asogun, Timothy D McHugh, Francine Ntoumi, Alimuddin Zumla and Richard Kock acknowledge support from the Pan-African Network for Rapid Research, Response and Preparedness for Infectious Diseases Epidemics – PANDORA-ID-NET, funded through the European and Developing Countries Clinical Trials Partnership (EDCTP) (grant number RIA2016E-1609). David Simons is supported by a PhD studentship from the UK Biotechnology and Biological Sciences Research Council (BB/M009513/1).

